# Machine learning-based forecasting of daily acute ischemic stroke admissions using weather data

**DOI:** 10.1101/2024.07.03.24309252

**Authors:** Nandhini Santhanam, Hee E. Kim, David Rügamer, Andreas Bender, Stefan Muthers, Chang Gyu Cho, Angelika Alonso, Kristina Szabo, Franz-Simon Centner, Holger Wenz, Thomas Ganslandt, Michael Platten, Christoph Groden, Michael Neumaier, Fabian Siegel, Máté E. Maros

## Abstract

**Background:** In the midst of the emerging climate crisis, healthcare providers lack locally validated, disease-specific surveillance models. Stroke, a significant contributor to the global disease burden, has been linked to climate change. Therefore, we developed and benchmarked machine learning (ML) models based on locoregional weather systems to forecast the number of daily acute ischemic stroke (AIS) admissions.

**Methods:** AIS patients diagnosed between 2015 and 2021 at the tertiary University Medical Center (UMC) Mannheim, Germany were extracted from the local data integration center and geospatially matched to weather data from the German Weather Service (DWD) based on the clinic’s, patients’ home and closest tower’s locations at the time of admission. Statistical- (Poisson), boosted generalized additive model (GAM), support vector machines (SVR), and tree-based models including random forest (RF) and extreme gradient boosting (XGB) were evaluated in regression settings within time-stratified nested cross-validation setup (training-validation: 2015-2020, test set: 2021) to predict the number of daily AIS admissions.

**Findings:** The cohort included 7,914 AIS patients (4,244 male, 53·6%). XGB showed the best test performance with lowest mean absolute error (MAE) of 1·21 cases/day. Maximum air pressure was identified as the top predictive variable. Shapley additive explanations analyses revealed that temperature extremes of extended cold-(lag-3 minimum temperature <-2 °C; minimum perceived temperature <-1·4 °C) and hot stressors (lag-7 minimum temperature >15 °C), as well as stormy conditions (lag-1 and lag-2 maximum wind gust >14 m/s and speed >10·4 m/s), increased stroke incidences substantially with distinct seasonal associations.

**Interpretation:** ML models can sufficiently forecast AIS admissions based on weather patterns allowing for improved resource allocation and preparedness.

## Introduction

The intensifying climate crisis poses a severe threat to ecosystems and human well-being, particularly to aging populations.^1–3^ Stroke is a major contributor to the global burden of cardiovascular disease, requiring prompt treatment for effectiveness.^1^ However, current healthcare systems struggle to dynamically adapt to weather related fluctuations in demand.^2,3^ This study leverages machine learning (ML) to develop predictive models using meteorological data to forecast acute ischemic stroke (AIS) admissions, aiming to enhance healthcare planning and accelerate responses to weather-related health incidents.

In addition to individual risk factors, various weather conditions have been linked to stroke occurrences, including extremes of ambient temperature,^4–8^ atmospheric pressure,^9–11^ wind speed,^12,13^ and ambient particulate matter with a diameter of <2.5 μm (PM_2.5_) pollution.^14,15^ Nonetheless, the results from these investigations have been inconclusive. Some studies have found a positive association between higher temperatures due to heat stress,^7,8^ higher air pressure,^10,11^ and higher wind speed^12^ leading to an increase in stroke occurrences. In contrast, other studies have established a negative link between AIS admissions and cooler temperatures,^4–6^ lower air pressure,^9^ as well as lower wind speed.^13^ While certain studies found no relevant association between weather conditions and the occurrence of stroke.^7,16^ Although ML models have been previously used to predict the number of admission counts for various diseases based on weather data, such as heat strokes, cerebrovascular, and overall emergency room visits^17,18^, none of these studies concerned with ischemic stroke admissions. Furthermore, they were neither intended for forecasting nor did they fully exploit the extensive array of weather features and lagged parameters to develop an open-source comprehensive predictive framework.

Therefore, this study aimed to develop and benchmark ML-based predictive models for AIS admissions using geospatially matched locoregional weather parameters for the clinically relevant daily time resolution. We employed a time-stratified 5x5-fold nested cross-validation setup over a seven-year period for a tertiary university clinic with a catchment area of 600,000 population. Our results underscore the potential of ML algorithms to forecast AIS admissions based on weather patterns, enabling improved resource allocation in the midst of climate change and providing a generalizable open-source framework applicable to various diseases.

## Materials and Methods

### Patient selection

This single-center retrospective cohort study entitled “Weather-based Stroke event and Outcome Risk Modelling (WE-STORM)” was approved by the local use- and access- (UAC) and ethics committees (Medical Ethics Commission II, Medical Faculty Mannheim, Heidelberg University, approval nr.: 2022-800R-MA). All methods were carried out following institutional guidelines and regulations. The ethics committee waived written informed consent due to the retrospective nature of the analyses. All patients admitted with suspected acute ischemic stroke between 2015-01-01 and 2021-12-31 at the University Medical Center (UMC) Mannheim, Germany, were retrieved from the local data integration center (DIC) using the core data set of the Medical Informatics Initiative (MII), which was based on standardized Health Level Seven International Fast Health Interoperability Resources (HL7 FHIR) specifications.^19^ Patients were identified using the International Classification of Diseases, Tenth Revision, German Modification (ICD-10-GM) codes: I63.0-9. Besides hospital diagnoses, general demographic information such as age, sex, admission date and patients’ home address (postal codes) were also extracted.

Postal codes were used to link local weather patterns to the admission date and time. In case of missing values, it was replaced with that of the clinic’s location (postal code). This information was then used to perform weather data extraction and geospatial matching for downstream analyses. If patients had multiple visits to the emergency department or outpatient ambulance, only the inpatient visit to the hospital was considered.

### Weather data

Weather data were retrieved from the open data server of the German Weather Service (DWD) using the *rdwd* package. It comprised 440 weather stations covering Germany. The stations measured various parameters, such as air temperature, relative humidity, pressure, wind speed, wind direction, sunshine duration, precipitation, and cloud cover, with different parameters being sampled at different temporal resolutions between 2015 and 2021. Because not all stations had all measurements, weather parameters with hourly and daily resolutions were collected from stations with full parameter coverage and based on the patients’ home locations (135/440, 30·7%) to optimize the balance between sufficiently detailed temporal resolution and data processing requirements (**Supplementary Fig. S1**). Weather features averaged over multiple days were represented as lagged variables, such as a 3-day average indicated by lag-3 or a 7-day average indicated by lag-7.

Additionally, we calculated well-established human biometeorological parameters such as perceived temperature (PT).^20^ PT is an index that jointly considers factors like air temperature, humidity, wind velocity, and radiation fluxes to quantify human thermal perception. It is derived from the Klima-Michael model, which uses an energy balance approach to describe the complex interactions of meteorological components on the body’s thermal equilibrium (German Weather Service, Glossary: K-Climate-Micheal model). PT was defined as the equivalent air temperature in an outdoor environment for a male reference subject (35 years, 1·75 m, 75 kg) with an internal heat production of 135 W/m^2^ (walking at 4 km/h on flat ground) in specific conditions (50% of relative humidity) and a reduced wind velocity (slight breeze).^20^ It is assumed that clothing is adapted to achieve thermal comfort. PTs between 0 and 20 °C mean comfort, <0 °C create a cold, and >20 °C a warm feeling, respectively.

### Geospatial matching

We assumed that patients were either at home or near their homes when the ictus occurred. To counteract this potential dependency, we performed a geospatial matching by assigning each patient to two weather stations, one closest to their home location and one closest to the clinic’s (UMC) location. This approach allowed us to identify three different patient types (**Supplementary Fig. S1**): 1) those with weather stations matched to their home location that were not the closest station to UMC 2) those that had the same station closely matched (<20km) both to their home and the clinic’s location, 3) like 2) but this station was situated further away (>20 km). Weather features were extracted from these respective stations for the admission date and time, and the previous seven days (lag-1 to lag-7) and were transformed into daily, two-day, weekly, and monthly resolutions (**Supplementary Table S1**). To adjust for seasonal- and long-term trends, we have also incorporated calendar-based variables such as weekdays or -ends, year, week number and holiday indicators based on public and school holidays in the state of Baden-Wuerttemberg as additional features in the feature space.

### Machine learning setup

The entire ML workflow was implemented using the *caret* package in the open-source R language (v.4.1.1, R Core Team, Vienna, Austria). All ML and statistical analyses were reported in accordance with the recently updated guidance for reporting clinical prediction models that use regression or machine learning methods (Transparent Reporting of a multivariable prediction model for Individual Prognosis Or Diagnosis, TRIPOD+AI statement).^21^ We investigated well-established shallow ML algorithms, including support vector regressors (SVR) using linear kernel (*e1071* package) and tree-based models including random forest (RF; *randomForest* package) and extreme gradient boosting (XGB; *xgboost* package) to predict the number of ischemic cases for the respective time resolution in a regression setting. Both ML- and statistical models were comprehensively evaluated within the same time-stratified 5x5-fold nested cross-validation (CV) setup with a training-validation set ranging from 2015 to 2020 and 2021 serving as the hold-out test set. (**Fig. 1**). In each fold, an additional year was incrementally added to the training set (sliding window approach). For consistency, each year was standardized to 365 days by excluding the leap year days (i.e. 2016, 2020; n=2). Weather input variables utilized in all the models underwent standardization through scaling and centring. Hyperparameters were tuned using predefined set of parameters within a grid search for the respective ML models. The root mean square error (RMSE) was used as loss function. All details regarding the training-validation setup and hyperparameter settings are available in the companion GitHub repository of the paper (https://github.com/MIDorAI/Machine-learning-based-forecasting-of-acute-ischemic-stroke-admissions-using-weather-data). Performance metrics of the tuned models reported on the test set (2021) were the RMSE, median absolute error (MAE), and mean absolute percentage error (MAPE). Feature importance rankings were calculated for RF using the built-in *VarImp* function of the *caret* package with the robust permutation-based variable importance setting^22^ and for XGB using gain-based importance. Additionally, SHapley Additive exPlanations^23^ (SHAP) values were calculated and plotted using the *kernelshap* package to aid visual interpretation of the results.

**Figure 1.**
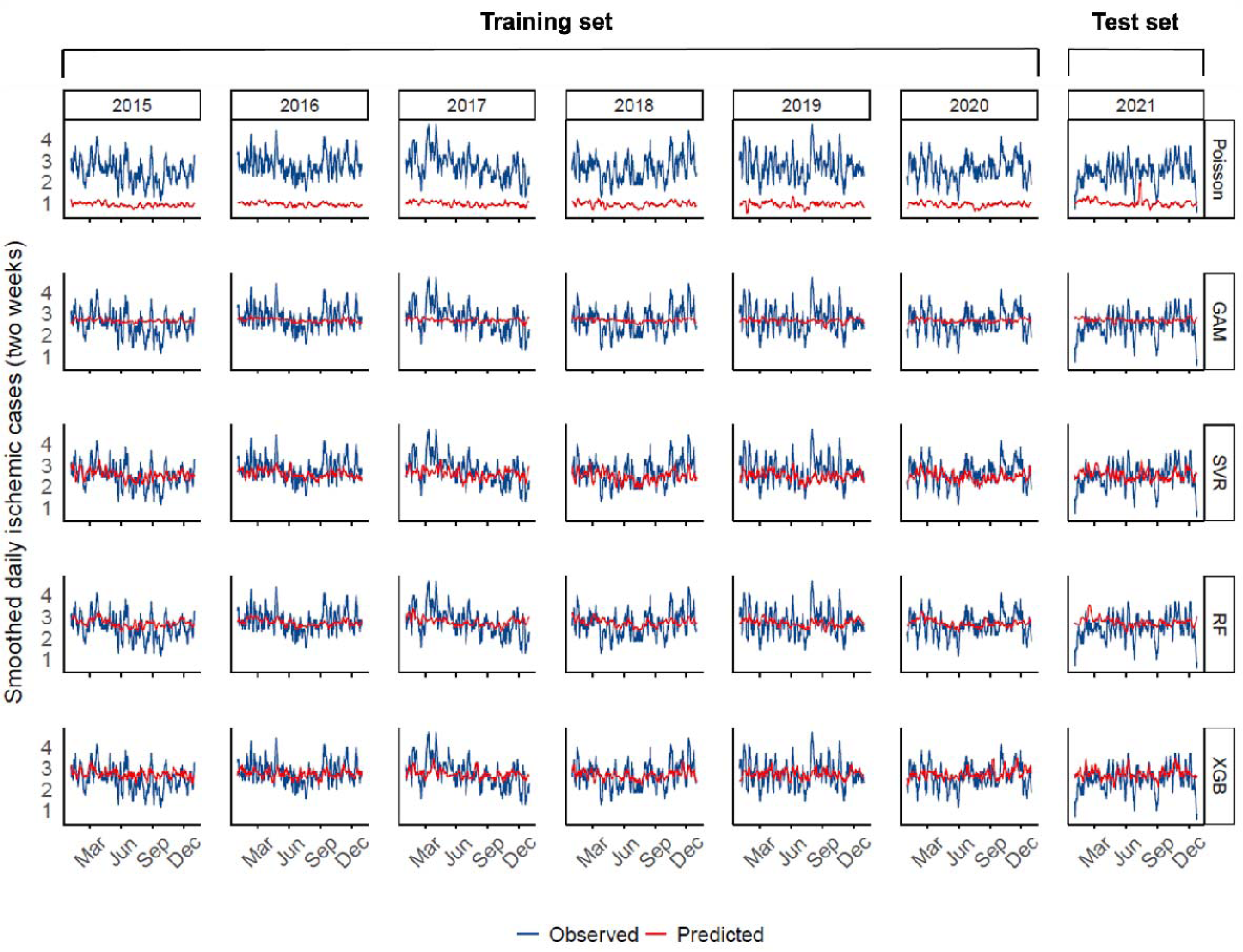
Setup for developing and benchmarking machine learning (ML) models to predict daily ischemic stroke admissions. Six years (2015 to 2020; n=2190 days) constituted the training set, wherein 5x5-fold, time-stratified, nested cross-validation was performed to optimize hyperparameters of the benchmarked ML models. The optimized models were then applied to the hold-out test set (2021; n=365 days) in a regression setting. The investigated ML models (horizontal facet panels) included both well-established statistical models like Poisson regression (baseline) and boosted generalized additive models (GAM) as well as shallow ML algorithms such as support vector regression (SVR), random forest (RF) and extreme gradient boosting (XGB). For each year (vertical facet panels), the daily number of observed (blue lines) and ML-predicted (red lines) AIS cases were smoothed for a two-week period.

### Statistical baseline models and analyses

As baseline statistical models, Poisson regression with a log-link distribution and boosted generalized additive models (GAM; *mboost* package) with a negative binomial distribution were fitted with the same set of input features and evaluated the same metrics (MAE, MAPE, RMSE) as the previously described ML models within R (v.4.1.1). However, the time-stratified nested cross-validation setup was applied only for GAMs. For this, the selection of specific basis functions for each variable was determined by the count of unique values present within them. We used the default (reduction score) variable importance for GAM provided in the *mboost* package. Reduction score quantifies the individual contribution to risk reduction of each base-learner and can thus be used to compare the importance of different variables in the model. Higher scores [0-1] indicate a greater impact on error reduction. The autocorrelation-(ACF) and partial autocorrelation function (PACF) plots of daily AIS cases were also evaluated to quantify longitudinal dependencies in the data (**Supplementary Fig. S2a & S2b**). Normally distributed variables were summarized as mean and standard deviation (SD), while non-normally distributed features were described with their median and interquartile range (IQR). Categorical variables were reported as proportions. Statistical significance was defined as two-sided p<0.05 and p-values were provided with their 95% confidence intervals (CI). Due to the explorative nature of our study, we did not adjust for multiple testing.^24^ Figures were created using the *ggplot2* and *leaflet* R packages using colour-blind safe palettes.

## Results

### Study cohort

A single-center retrospective cohort of 7914 (4244 male, 53·6%) patients admitted with AIS between 2015 and 2021 at the UMC Mannheim, Germany was retrieved from the local DIC. The average age of patients was 71 years (range: 7-98 years, SD=14 years). The descriptive statistics for stroke admissions in the cohort and the distribution of the weather parameters such as temperature [°C], relative humidity [%], pressure [hPa], and windspeed [m/s] in the feature space (overall n=133 variables) were summarized in **Table 1**.

**Table 1.**
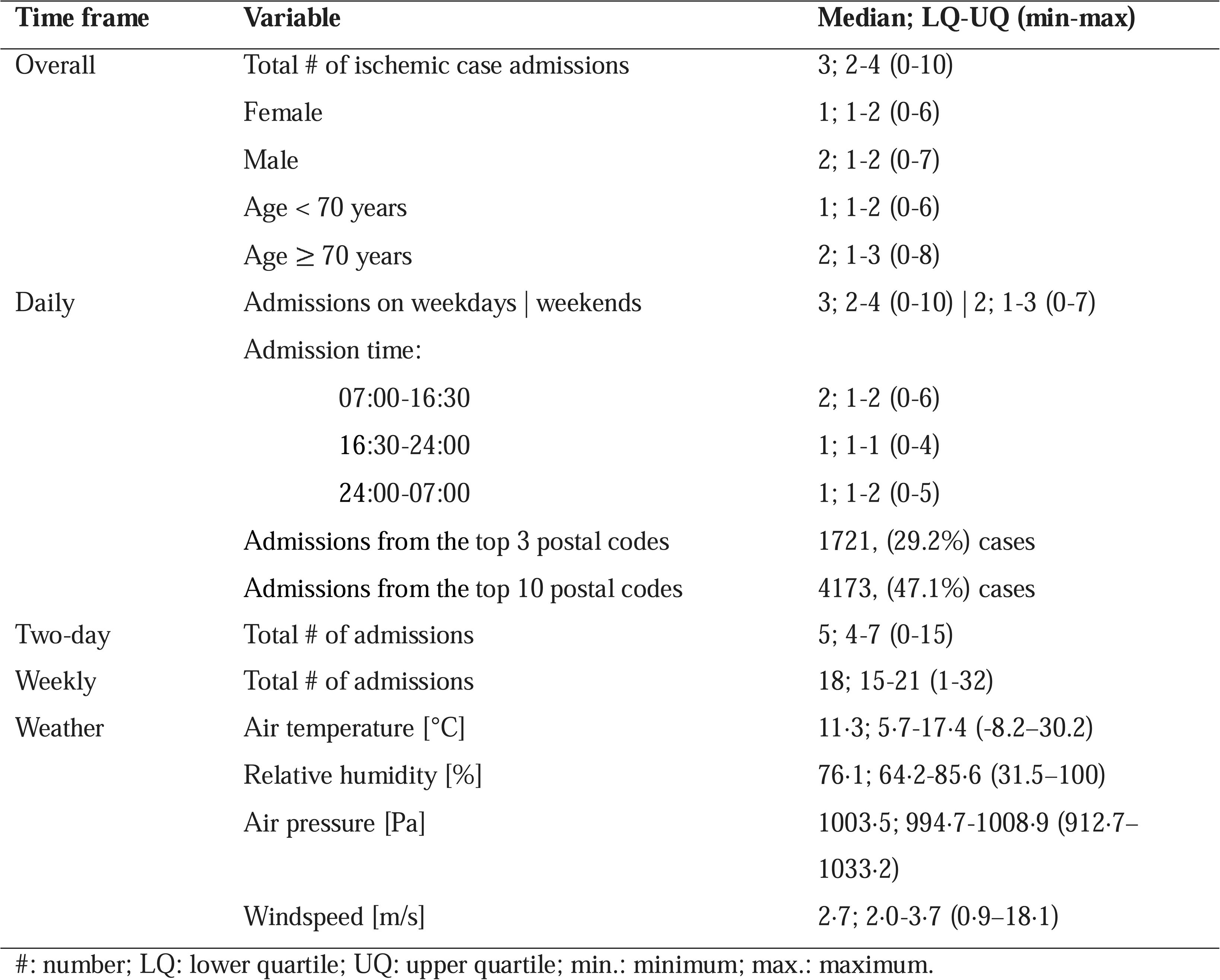
Summary table of the study cohort including the distributions of ischemic stroke admissions and weather parameters.

The ACF and PACF plots (**Supplementary Fig. S2a & S2b**) of daily AIS cases showed no significant autocorrelation (lags 1-3: ACF: p_lag1_= 0·31; p_lag2_= 0·06; p_lag3_= 0·071; PACF: p_lag1_= 0·084; p_lag2_= 0·093; p_lag3_= 0·12). This indicated that time series data could be considered sufficiently stationary, therefore, suggesting the applicability of classical shallow ML models.

### Yearly and seasonal trends

Yearly trends in AIS admissions displayed a significant increase in 2015 with subsequent declines in 2018 and 2020 (**Fig. 2a**). Analysis of aggregated monthly data over the seven-year period revealed pronounced seasonal variations with peak incidences occurring in March (mean=88·85, 95% CI: 75·47-102·24, p=3·46X10^-6^) and a decrease in September (mean=74·71, 95% CI: 62·64-86·78, p=5·21X10^-6^, **Fig. 2b**) followed by a secondary peak in October (mean= 84·57, 95% CI: 76·65-92·48, p=2·06X10^-7^) and November (mean=83·71, 95% CI: 77·18-90·24, p=6·89X10^-6^). In contrast, if the case count was averaged over the week (week 1 to 52) over the 7-year period, no consistent pattern could be observed (**Fig. 2c**) other than noticeable dips during the holiday season (50^th^-2^nd^ weeks).

**Figure 2.**
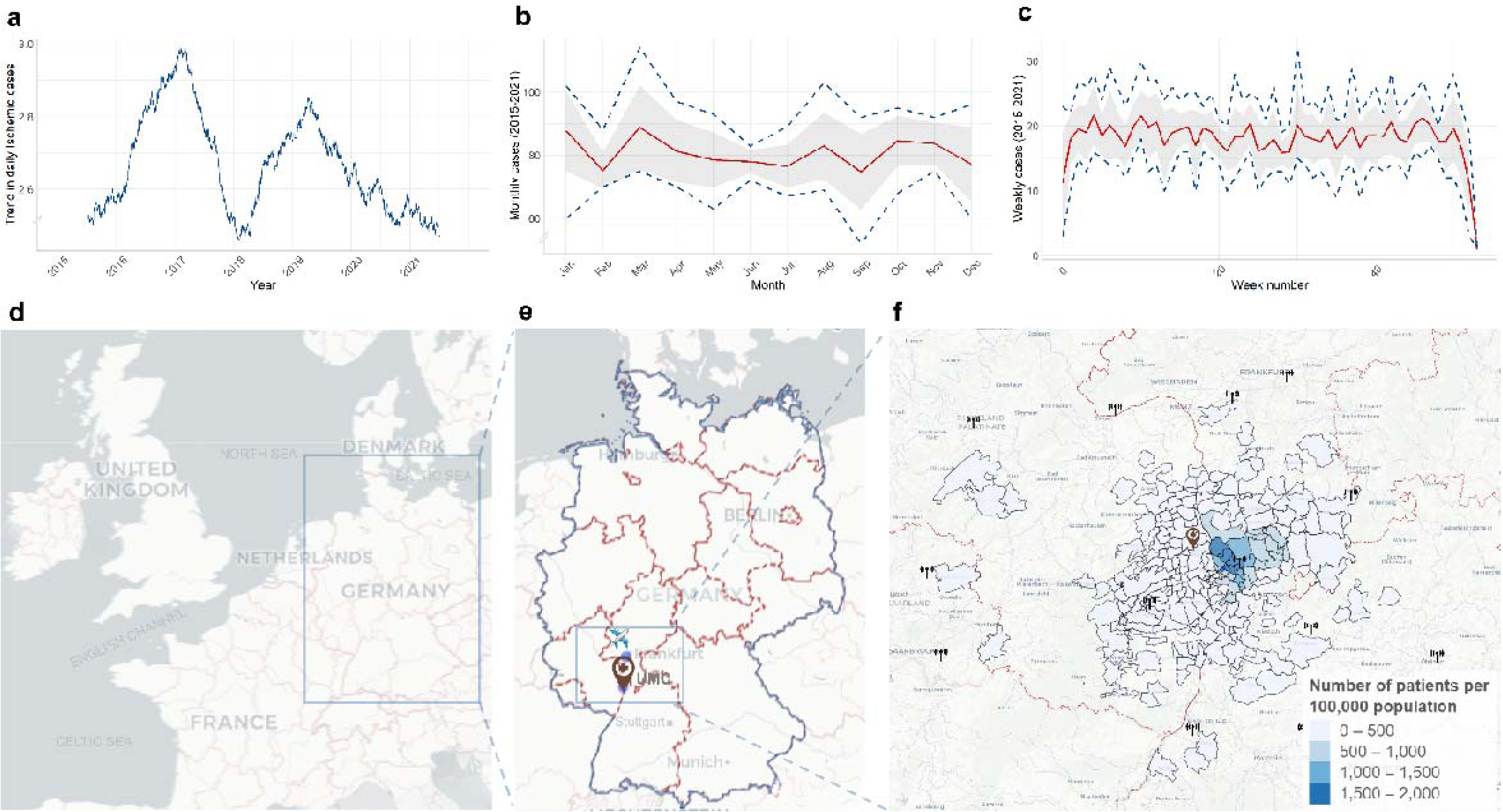
Combination figure of yearly, monthly and weekly acute ischemic stroke admission (AIS) and their geospatial distribution. (**a**) Yearly trend analysis of AIS case counts showed a pronounced increase from 2015 to 2017, resembling hype cycles, potentially attributable to landmark clinical trials for the endovascular treatment of AIS.^31^ In contrast, during the early COVID-19 pandemic (2020-2021), a clearly decreasing trend was observed. (**b**) Monthly AIS admissions (averaged over the 7-year study period; red line) indicated seasonal peaks in March, October, and November (95% CI in shaded gray) with min-max ranges (dark blue dashed lines). (**c**) Weekly averages showed no apparent trends except for noticeable dips during the holiday season (50^th^-2^nd^ weeks). (**d**) The University Medical Center Mannheim, Germany (UMC; 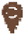 is located in the state of Baden-Wuerttemberg at (**e**) the corner of a German tri-state area (Rhineland Palatinate and Hesse; light blue bounding box). UMC is the primary tertiary care provider in Mannheim, the largest city of the region and the second largest in the state, with a population of 310,000 and a catchment area of over 600,000 people between Frankfurt ( ) and Stuttgart. (**f**) Geospatial distribution highlighting the density of ischemic strokes per 100,000 population in the catchment area of UMC using postal code-based distribution of patients’ home locations. The top three contributing areas were within an <11 km radius of the clinic and accounted for 29·2% of the total patient count, while 96·2% of all admissions arrived from a <50 km range. The selected weather stations are indicated with the icon (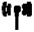).

### Spatial distributions

The UMC Mannheim is located in the state of Baden-Wuerttemberg (**Fig. 2d** & **2e**) in the largest city of the metropolitan region Rhine-Neckar. Postal code-based geospatial analyses of patients’ home locations showed that the top three contributing areas were within a <11 km radius of the clinic and contributed to 29·2% of the total patient count. Over the span of seven years, patient counts from these regions ranged between 400-600 admissions/year (**Fig. 2f**). Similarly, when assessing the prevalence of the condition on a standardized rate per 100,000 population, these three postal codes consistently emerged as the top contributors. Overall

∼96·2% of admissions came from within 50 km radius of the clinic supporting our locoregional approach and assumptions.

### Baseline statistical models

The baseline Poisson model estimated (**Table 2**) the lag-5 mean cloud cover (OR=0·97, 95% CI: 0·93-0·98, p=0·0032) and lag-1 mean pressure (P_mean_lag1_; OR=0·45, 95% CI: 0·24-0·81, p=0·0076) to be negatively correlated with AIS admissions. Conversely, lag-2 minimum temperature (T_min_lag2_; OR=1·10, 95% CI: 1·01-1·20, p=0·025) and maximum wind gust (V_max_; OR=1·02, 95% CI: 1·00–1.04, p=0·017) were positively associated with increased daily cases counts.

**Table 2.**
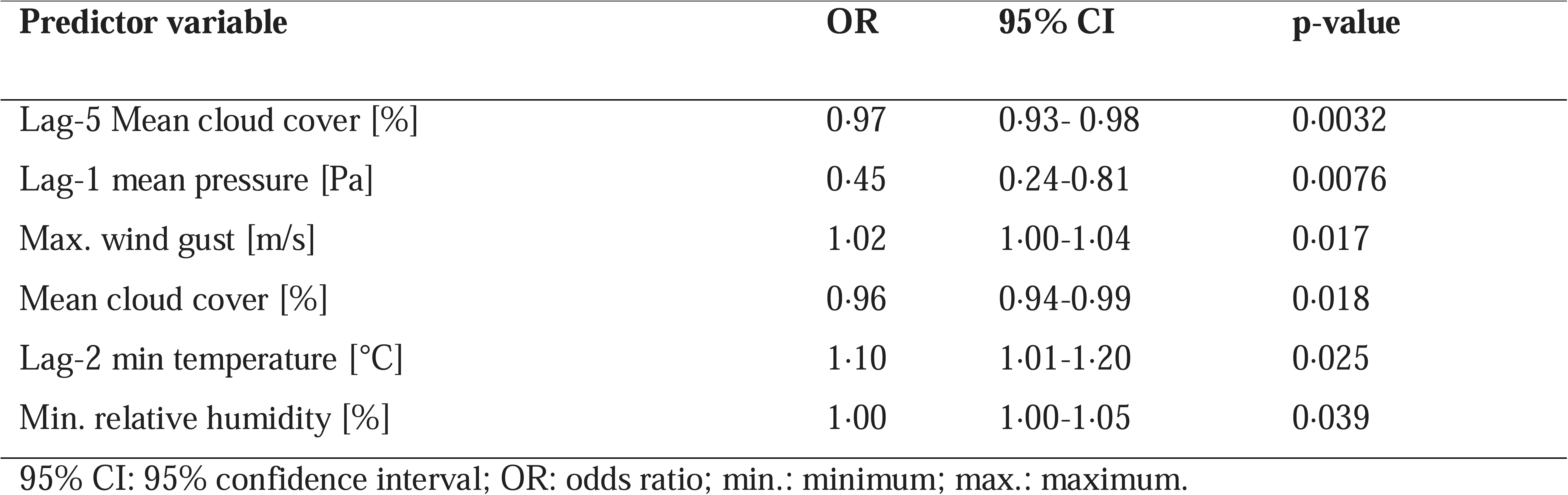
Summary table of the top six predictor estimates of the Poisson regression model.

The GAM identified maximum-(P_max_) and mean pressures (P_mean_) as the two most influential variables with a reduction score with respect to RMSE in percentages of 0·85 and 0·53, respectively (**Supplementary Fig. S3a**). The third most influential variable was found to be minimal PT (PT_min_) with a reduction score of 0·13. Weekends were selected as one of the top ten features.

### Machine learning models

Among the ML models tested, XGB demonstrated the highest performance in predicting daily AIS cases, with the lowest MAE of 1·21 cases/day and RMSE of 1·49 cases/day in the test set. This represented a reduction of ∼29% in MAE and a 44% decrease in RMSE compared to the baseline Poisson model (**Table 3**). Notably, similar performance was observed when the model was exclusively trained with weather variables only (**Table 3**).

**Table 3.** Overview of baseline statistical and ML model performance metrics on test set (2021).

| Model type | Model name | Weather features only |  |  | Weather and calendar features |  |  |
| --- | --- | --- | --- | --- | --- | --- | --- |
|  |  | MAE<br>(N<sub>count</sub>/day) | MAPE<br>(%) | RMSE<br>(N<sub>count</sub>/day) | MAE<br>(N<sub>count</sub>/day) | MAPE<br>(%) | RMSE<br>(N<sub>count</sub>/day) |
| Baseline | Poisson | 1.72 | 58 | 2.73 | 1.69 | 57 | 2.68 |
| statistical | GAM | 1.44 | 56 | 1.64 | 1.41 | 56 | 1.62 |
| Machine learning (ML) | SVR | 1.28 | 52 | 1.58 | 1.27 | 52 | 1.58 |
|  | RF | 1.26 | 49 | 1.54 | 1.25 | 49 | 1.52 |
|  | XGB | 1.25 | 48 | 1.52 | 1.21 | 47 | 1.49 |

GAM: generalized additive model; MAE: median absolute error (N<sub>count</sub>/day); MAPE: mean absolute percentage error (%); RMSE: root mean square error (N<sub>count</sub>/day); RF: random forest; SVR: support vector regression using linear kernel; XGB: extreme gradient boosting.

The XGB model effectively captured the variability of daily AIS case counts, especially for counts between 2 and 5 in the hold-out test set (**Fig. 1**). However, the model encountered limitations when predicting days with either extremely low (0, 30/365, ∼8·5%) or high (>5, 12/365, ∼3·3%) case counts in the test set (2021). This performance pattern mirrored the distribution of the training set from 2015-2020 wherein low (0, 147/2190, ∼6·7%) and high (>5, 129/2190, ∼5·9%) AIS admissions were also infrequent. Additionally, 67.4% of the cases in the hold-out test set and 69·4% of the training dataset cases fell within the 2-5 case count range.

### XGB-based variable importance of weather parameters

P_max_ consistently emerged as the top variable for forecasting daily AIS case admissions across all deployed ML models. The XGB model distinguished itself by identifying lag-3 minimum temperature (T_min_lag3_) as the second most relevant variable (**Fig. 3a**); while SVR and RF models selected mean pressure (P_mean_) for this position (**Supplementary Fig. S3b & S3c**). Interestingly, PT_min_ emerged as the third-ranked predictor in both the XGB and RF models.

**Figure 3.**
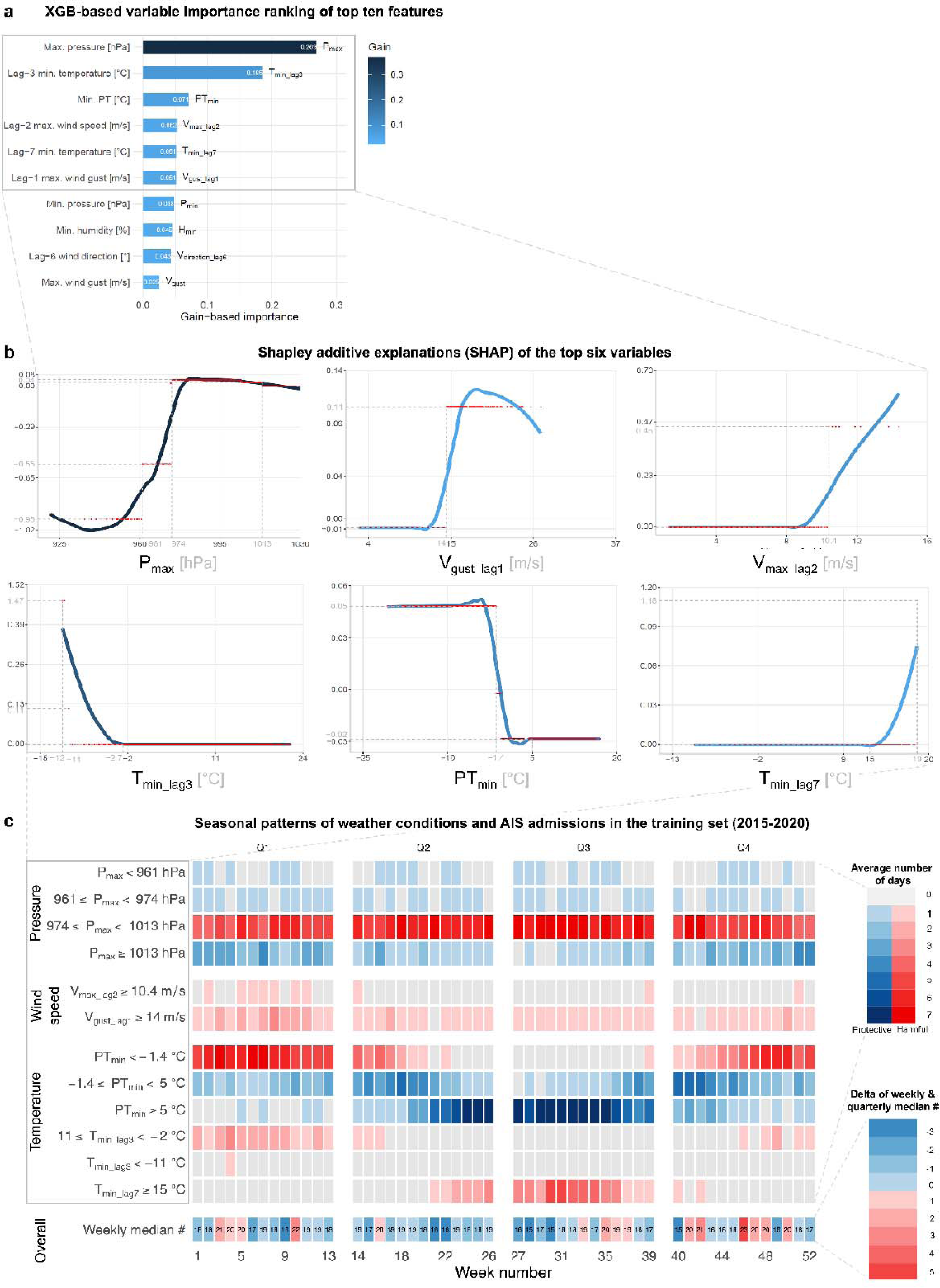
Composite figure of detailed analyses of the most important predictors of the best performing XGB model and their link to seasonal distribution of daily AIS admissions. (a) Horizontal bar charts of the top ten most relevant features using normalized gain-based variable importance ranking of the best XGB model. **(b)** Shapley additive explanations (SHAP) of the top six variables, including (upper-row) maximal air pressure (P ), lagged 1- and 2-days maximal wind speed (V ) and wind gust speeds (V ); and (lower-row) minimal lagged 3-days temperature (T_min_lag3_), minimal perceived temperature (PT_min_), and 7-days minimal temperature (T_min_lag7_). These variables (light grey bounding boxes) accounted for an overall sum of 0.84 gain-based importance out of the 133 investigated weather and calendar features. Inflection points on the subplots indicate (grey dashed lines) when the respective variable’s effect was associated with an increase or decrease in AIS counts**. (c)** Heatmaps indicating the seasonal distributions of specific weather conditions in the training data (2015-2020; n=2190 days), thresholded using respective values from SHAP inflection points **(b)**; dashed grey lines and bounding box), and faceted as pressure, wind, and temperature features. The number of days that the respective condition has occurred was calculated by jointly aggregating at yearly and weekly levels (matrix: 7 [years] x 52 [weeks] rows by 12 columns [respective conditions]). This matrix was then reduced to the weekly level to calculate the median number of days these conditions occurred across different years (matrix: 52 [week-median] rows by 12 columns [respective conditions]). These protective (shades of blue) or harmful (red) median number of days were then color-coded based on the sign of the SHAP values. Additionally, the deltas of weekly AIS case counts (aggregated over the seven years) were compared against the respective quarterly medians (ordinal color key, lower-right corner: -3 to +6, from blue to red) and displayed with exact count values in the “overall” facet. P_max_ showed a sigmoid-like link as (very) low pressures (P_max_<960) substantially decreased stroke admissions (SHAP=-0·95), while medium-high values (974-1013 hPa) were associated with an increased stroke incidence all year round (Q1-Q4). High-pressure (P_max_≥1013 hPa) conditions during winter (Q1, Q4) occurred less frequently and were only marginally protective. Also linked to high-pressure, cold stressor days (Q1, Q2, and Q4) and associated windy conditions (V_max_lag2_>10·4 m/s and V_gust_max_lag1_>14 m/s) substantially increased AIS admissions (SHAP_Vmax_=0·11 and SHAP_Vgust_=0·45). Similarly, extended cold stressor periods during winter with T_min_lag3_<-2 °C or PT_min_<-1·4°C were strongly linked to more strokes (SHAP up to 1·47). Conversely, PT_min_ in classical temperate ranges (-1·4<PT_min_<20°C) were slightly protective (SHAP=-0·03), although these effects could be outweighed (SHAP up to 1·18) during extended heat stress periods (T_min_lag7_>15°C) of the summer (Q3).

The top 10 most important variables for XGB focused on temperature- and wind speed-related features, while RF emphasized temperature- and vapor pressure-based features. Besides weather variables, weekends also emerged among the top ten variables in the XGB model.

The SHAP-based analyses provided insights into how the top six meteorological parameters of XGB influenced the prediction of daily AIS admissions (**Fig. 3b**). P_max_ values were categorized into four ranges in post hoc analyses for a more detailed interpretation. During low pressure conditions (<961 hPa), the SHAP was -0·95, indicating a reduction in AIS cases (**Fig. 3b**), which occurred on ∼1·4% of the training days (31/2190) over a span of 24 weeks (**Fig. 3c**). The second range (961-974 hPa), also exhibited a protective effect, reducing ischemic cases by -0·55, despite being observed on only 2·5% of the days (56/2190). Conversely, high-pressure (ranging 974-1013 hPa) conditions were positively associated with an increase in AIS case counts (SHAP=0·04), occurring on 76% (1664/2190) of the training days without seasonal preference (**Fig. 3c**).

Temperature extremes played a dual role on AIS admissions, with both cold and hot stressor days positively associated with higher ischemic stroke counts. Especially, prolonged cold stressor periods with T_min_lag3_<-11 °C were been strongly linked (SHAP=1·47) with a surge in AIS admissions (**Fig. 3b**). While such weather conditions were observed on only five days in the entire dataset, stroke admissions peaked between 7-9 cases exclusively on these days, specifically during January in the years 2017 and -18 (**Fig. 3c**), however, AIS counts were generally lower on days surrounding these extreme cold periods. Additionally, PT_min_<-1·4°C underscored the positive impact of cold stress with an increased SHAP value of 0.05 (**Fig. 3b**). Such conditions were prevalent during the winter months (in the 1^st^ and 4^th^ quarters) and affected 36% (788/2190) of the training days (**Fig. 3c**). Similarly, prolonged hot stressors were also identified as triggers that increased ischemic stroke occurrences, with the 7-day lagged Tmin (T_min_lag7_) ranked as the 5^th^ most important variable by XGB (**Fig 3a**). When T_min_lag7_ exceeded 15 °C (256/2190, 11·6%), the SHAP values steeply increased in a quasi-linear fashion from 0 to ∼0·08 **(XSFig. 3b)**. If T_min_lag7_ reached 19 °C, the SHAP value escalated punctually to 1·18 (**Fig. 3b**). This phenomenon had a seasonal preference during the 3^rd^ quarters, especially during the months of August and September in the years 2018-2020 (**Fig. 3c**).

Wind-related variables exhibited a distinct pattern indicating short-term effects on AIS admissions, particularly for lag-2 maximum wind speed (V_max_lag2_) and lag-1 maximum wind gust (V_gust_lag1_), which were associated with an increase in AIS cases (SHAP=0·45 at V_max_lag2_=10·4 m/s and SAHP=0·11 at V_gust_lag1_=14 m/s; **Fig. 3b**). These windy conditions occurred primarily during the first quarter, aligning with periods of cold stress and high-pressure stormy conditions (**Fig. 3c**).

## Discussion

We developed and benchmarked a set of well-established ML and statistical models to investigate the association between locoregional weather systems and the number of AIS admissions over a seven-year period to enable better planning of clinical resources. We found that shallow ML models were sufficient and outperformed baseline statistical models by 20-40% in terms of MAE and RMSE. XGB performed the best with an average MAE of 1·21- and RMSE of 1.49 cases/day, making it potentially useful for real-time forecasting.

Regarding weather conditions, both cold and hot stressors days increased the number of daily stroke admissions, with prolonged colder conditions having a more prominent effect. Additionally, high pressure and stormy conditions tended to increase daily AIS admissions. Our results highlight the potential application of ML models to forecast stroke occurrences based on weather- and seasonal patterns in real-time for optimal clinical resource allocation and patient care. Furthermore, our time-stratified, nested cross-validation setup provides a general framework that can be used for various diseases in both single- or multi-center applications.

We observed a dual impact of temperature as both cold and hot stressors, especially over multiple days (>3 or 7 days), were associated with an increase in AIS counts, with a slight predominant effect of cold stress. This was consistent with findings from previous studies across diverse climatic zones globally.^4–6,25–27^ A retrospective analysis of hospital data in the United States revealed a surge in stroke admissions during winter, accompanied by increased mortality rates.^4^ Ambulance dispatches for ischemic stroke cases in Japan exhibited a similar seasonal pattern and were observed to be more common during lower temperatures.^5^ Likewise, a retrospective study in China over a two-year period reported that 1·57% of ischemic strokes could be attributed to extreme cold temperatures, particularly in the 0-7 day lag period.^6^ This aligns with our findings that prolonged cold stressors with T_min_<-11 °C on three consecutive previous days substantially increased stroke incidence. Furthermore, both tree-based models (RF and XGB) identified PT_min_ as one of the top three predictors of daily AIS admissions, thereby emphasizing the importance of key human biometeorological features. PT_min_ showed a bimodal distribution of the estimated effects with higher weights given for cold days (PT_min_<-1·4°C).

Heat stress has also been observed to increase the incidence of AIS.^7,8,28^ In a single-center retrospective study in Korea, Han et al. found that the seasonal AIS incidence in summer was significantly higher than in winter, and the mean temperature was positively associated with ischemic stroke with an RR of 1·006.^7^ In accordance with this, the baseline Poisson model in our study identified a positive association between the minimum lag-2 temperature and daily stroke cases, with an increase of ∼11% for every 1 °C increase. Ma et al. utilized a universal thermal climate index to quantify the weather conditions in Beijing^8^ and identified that the risk of suffering an AIS increased with heat stress, especially in the 45-65 years age group.

Multiple studies have shown that fluctuations in atmospheric pressure and temperature can promote arterial blood pressure instability and hemodynamic changes in the circulatory system.^12,26^ The association between atmospheric pressure and the incidence of stroke has been studied in multiple retrospective studies.^9–11^ Jimenez et al. reported that the drop in atmospheric pressure compared to the previous day can largely explain seasonal and daily variations of stroke incidence.^10^ Qi et al. also discovered that mean, minimum and maximum barometric pressures showed statistically significant positive associations with ischemic stroke occurrences and the colder season tended to be the more risk-prone.^11^ Our study also strongly supports these findings, as all ML models and the GAM identified pressure-related variables as the primary predictor of daily AIS admissions.

Coupled with high-pressure, stormy phenomena, maximum wind speed, and wind gusts on previous two days were linked to an increased number of AIS cases in our cohort. Similarly, in a small, localized study comprised of 409 stroke patients admitted during a two-year period (2006-2007) on an island in South Korea, wind speed and wind chill index were identified to have positive associations with AIS cases, which was more pronounced in spring and winter.^12^ This cumulative effect of cold and stormy conditions within the same seasonal window was also observed in our study, which implied a compounded effect. It is important to note, however, that air pressure measurements by the DWD towers are referenced back to the respective sea level. For our West-German region, it meant either the North Sea or the Atlantic Ocean.

Most studies analysing the association between meteorological parameters and the onset of ischemic stroke have predominantly utilized classical statistical models, such as Poisson regression or its variations.^11,14,25^ Only few studies have employed ML-based models to develop a predictive framework using weather parameters, while these focused on conditions like heatstroke or general emergency room admissions.^17,29^ Ogata et al. developed predictive models to forecast heatstroke admissions for a three-year time period for sixteen cities in Japan using models such as GLM, GAM, and XGB.^17^ It is noteworthy that the number of daily heatstroke admissions in their study substantially outranged (up to 400+ cases/d) the daily AIS admissions in our cohort (0-10 cases/d). The conventional GAM model exceeded the performance of other models, registering the lowest RMSE of 2·47 cases on their test set (2018) while XGB achieved an RMSE of 3·28. In contrast, XGB showed the best performance with an RMSE of 1.49 on the hold-out test set (2021) of our study for daily AIS admissions, despite predicting sparse values with lower variance, which is expected to be technically more challenging.

A specialized feature selection approach was used by Tuominen et al. to select the most robust weather parameters to forecast the combined daily emergency department arrivals in Tampere, Finland.^29^ In their analyses, they considered 158 explanatory variables, which closely mirrors the 133 features incorporated into our daily forecasting model. Besides shallow ML algorithms, we also explored various deep learning architectures,^30^ including recurrent neural networks (RNN) and long short-term memory (LSTM) networks, and complex forecasting frameworks like NeuralProphet by Meta. However, the choice to use shallow ML models was justified by the missing higher-order autoregressive associations within the data set. This allowed us to opt for the less hardware-intensive CPU-bound modelling setup, not requiring GPUs, hence making our approach more widely applicable.

This study has certain limitations, as it was a single-centre retrospective analysis. However, the substantial cohort size of ∼8,000 patients with a catchment area of ∼600,000 individuals supports the reliability of our results.^5^ Additionally, we utilized a fine-grained temporospatial matching method to select weather variables from various DWD stations corresponding to patients’ home locations and admission hours. The assumption that patients were in reasonable vicinity of their home address had to be made for downstream analyses. Over 96·2% of the admitted patients’ homes were located within a 50 km radius, wherein variations of weather patterns are expected to be minimal, thereby supporting the feasibility of this approach.^27^ It is noteworthy that we did not apply additional feature selection methods,^29^ but utilized the internal variable selection provided by the respective ML model during training and validation.^17^ Despite the established association of air pollutants with cerebral and respiratory diseases in previous studies, ^14,15^ we could not include this data in our analyses due to the very low density of monitoring stations in the area. Based on findings of the global, regional, and national burden of stroke and its risk factors study by Feigin et al., weather information can explain ∼10% of the variance of AIS incidence.^1^ Regardless, we found that ML models can predict the number of daily admissions with an acceptable MAE and RMSE of <1·5 cases/day, particularly for the most common band of daily ischemic cases between 2-5, covering ∼70% of the investigated timeframe.

In conclusion, using a detailed temporal and geospatial matching technique, this study systematically compared baseline statistical and ML models to forecast the number of acute ischemic stroke admission based on weather patterns. ML models outperformed classical statistical models, demonstrating their potential for real-time healthcare resource allocation. The best-performing model (XGB) identified atmospheric pressure, lagged temperature, PT_min_, and wind speed as most important predictors of stroke occurrence. Our results further emphasize the dual role of temperature for both hot and cold stressor days and the crucial effect of prolonged stormy conditions. We developed a generalizable framework that can be applied to various diseases and easly deployed as multi-centric applications in data integration centers nationally and around the world to determine the impact of locoregional weather conditions and seasonal variations.

## Data sharing statement

The deidentified count data of acute ischemic stroke admissions during the seven-year study period, supporting the conclusions of this article, are available from the corresponding author upon reasonable request.

## Author contributions

M.E.M. conceived the study. N.S. conducted data extraction and weather data matching. N.S., M.E.M., D.R., and A.B. performed statistical and machine learning modeling and analysed the data. N.S. and M.E.M. created the figures and tables. N.S. and M.E.M. wrote the manuscript. A.B. and D.R. gave technical support and implemented additive-based models. S.M. provided weather features and advised human biometeorological aspects of the study. A.A., K.S., M.P., K.S., and C.G. provided data. H.K. performed data extraction. A.A., K.S., M.P., CG.C., FS.C., H.W., and M.E.M. advised clinical aspects of the study. M.N., T.G., and F.S. advised the study. M.E.M. acquired funding and supervised the study. All authors critically reviewed and approved the final manuscript version.

## Data Availability

The de-identified count data of acute ischemic stroke admissions during the seven years study period, supporting the conclusions of this article are available from the corresponding author upon reasonable request.

