## Supplementary Appendix for "Machine learning-based forecasting of daily acute ischemic stroke admissions using weather data"

#### Supplementary table

**Supplementary Table 1. Summary table of the different features that are utilized in the fitted statistical and machine learning models.**

| Type | Feature [Unit] | Variable conversion |
|---|---|---|
| Weather | Pressure [hPa] | Mean, min, max, and lagged 1-7 days |
| | Vapour pressure [hPa] | Mean, min, max, and lagged 1-7 days |
| | Temperature [°C] | Mean, min, max, and lagged 1-7 days |
| | Dewpoint temperature [°C] | Mean, min, max, and lagged 1-7 days |
| | Perceived temperature [°C] | Mean, min, max |
| | Humidity [%] | Mean, min, max, and lagged 1-7 days |
| | Humid temperature [°C] | Mean, min, max, and lagged 1-7 days |
| | Sunshine duration [h] | Total and lagged 1-7 days |
| | Precipitation height [m] | Mean, min, max, and lagged 1-7 days |
| | Cloud cover | Status signal |
| | Wind speed [m/s] | Mean, min, max, and lagged 1-7 days |
| | Wind gust [m/s] | Mean, min, max, and lagged 1-7 days |
| | Wind direction [°] | Mean and lagged 1-7 days |
| Calendar | Year | Status with values 2015 -2021 |
| | Weekday status | Weekday or weekend |
| | Week number | Status with values 1-52 |
| | Holiday status | Yes/No |

min.: minimum; max.: maximum; lagged 1-7 days: lag-1 to lag-7

### Supplementary figures

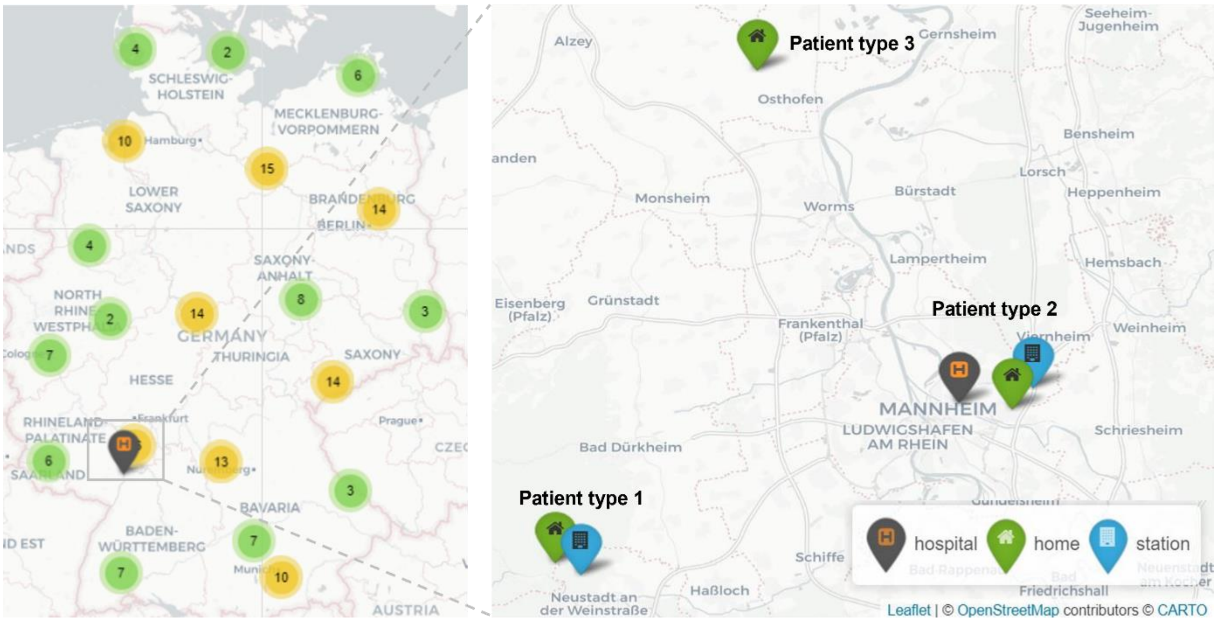


**Supplementary Figure S1. Distribution of German weather stations and patient types identified during geospatial matching.** Weather stations ($n_{station}$=135) that had full coverage of data for the seven years (2015-2021) of all selected variables ($n_{var}$=133) were chosen. When stations were close to each other, they were clustered together and shown as number of stations in that region in circles (number of towers <10: green; ≥10: yellow). The location of University Medical Center (UMC) was indicated with the gray pinpoint with orange H. Three different patient types were identified during the complex geospatial matching: 1) type 1 patients had a different measurement station (blue pinpoint) mapped to their home locations (green pinpoint) than the closest station to the UMC; 2) For type 2 cases, the same station was mapped as closest to both home and UMC locations (<20km away); 3) type 3 patients also matched to the same station, but their distance was >20 km to UMC and its reference station.

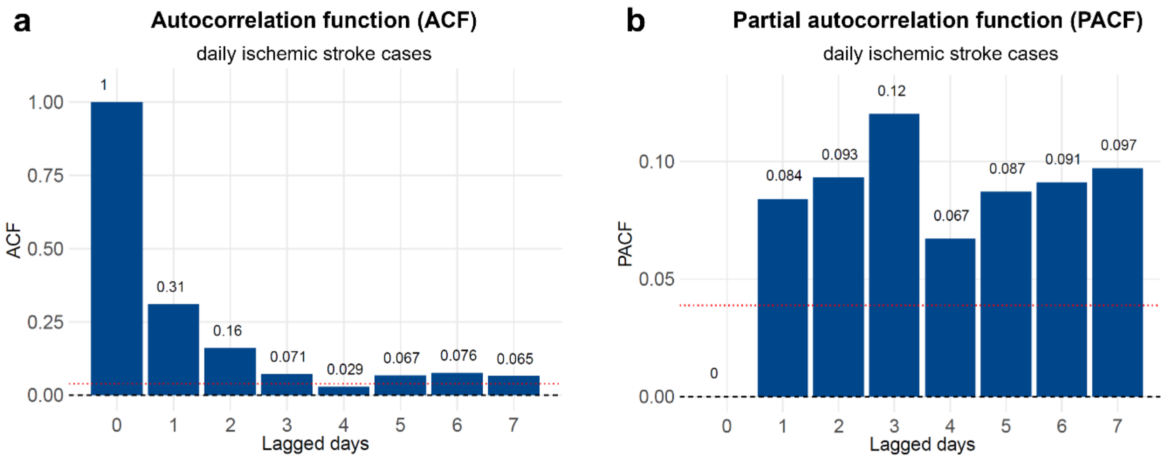


**Supplementary Figure S2. Autocorrelation- (ACF) and partial autocorrelation functions (PACF) plots of the daily ischemic stroke admissions.** The daily number of ischemic stroke admission were converted into time series, and the corresponding (**a**) ACF and (**b**) PACF with p values of the one- to seven-day lagged components were calculated and visualized as bar graphs. ACF p values were: $p_{lag-1}$=0.31, $p_{lag-2}$=0.16, $p_{lag-3}$=0.071, $p_{lag-4}$=0.029, $p_{lag-5}$=0.067, $p_{lag-6=}$0.076, and $p_{lag-7}$=0.065. PACF p values were: $p_{lag-1}$=0.084, $p_{lag-2}$=0.093, $p_{lag-3}$=0.12, $p_{lag-4}$=0.065, $p_{lag-5}$=0.087, $p_{lag-6}$=0.091, and $p_{lag-7}$=0.097. The ACF p value dropped considerably after lag-1, indicating that the influence of past number of admissions on current admissions quickly diminished after the one to two days. The PACF values did not have sharp cut-off and remained low, suggesting no strong autoregressive structure in the underlying data, making moving average or autoregressive components unnecessary for this analysis.[1] Consequently, shallow machine learning models were applied during downstream analyses.

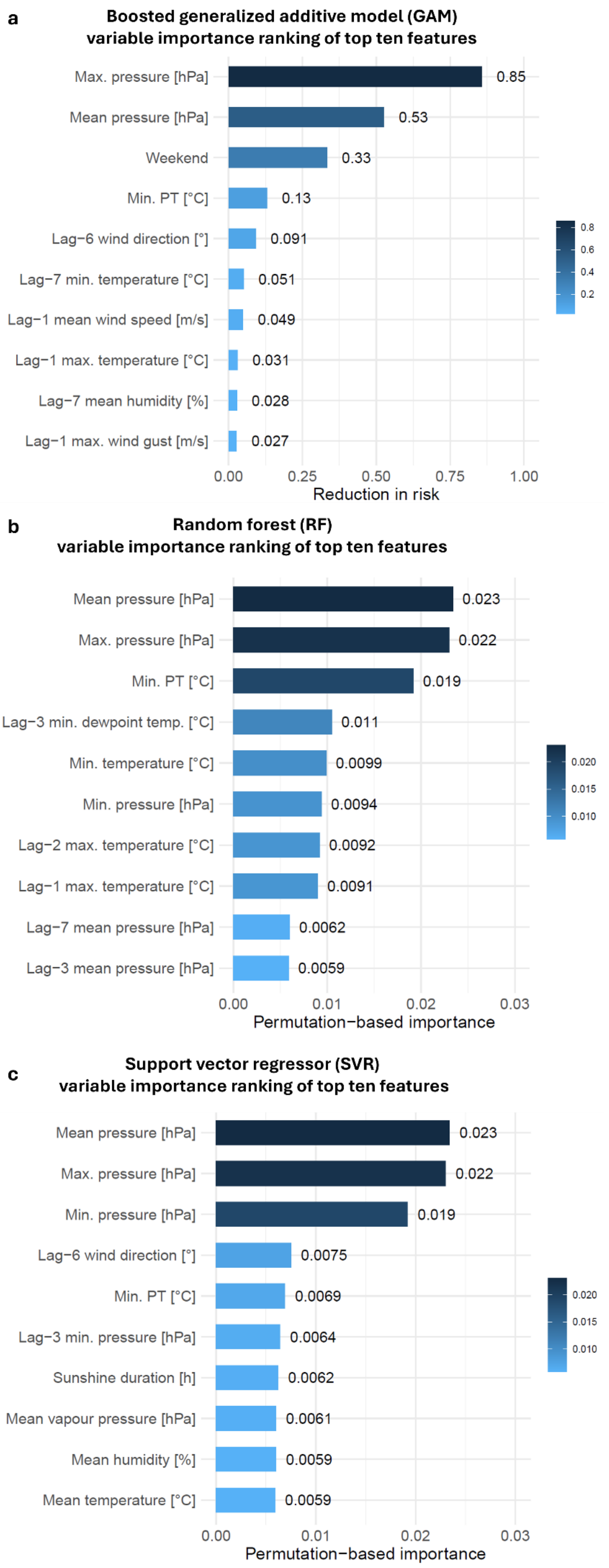

a
Boosted generalized additive model (GAM)
variable importance ranking of top ten features
Max. pressure [hPa]
0.85
Mean pressure [hPa]
0.53
Weekend
0.33
Min. PT [°C]
0.13
Lag−6 wind direction [°]
0.091
Lag−7 min. temperature [°C]
0.051
Lag−1 mean wind speed [m/s]
0.049
Lag−1 max. temperature [°C]
0.031
Lag−7 mean humidity [%]
0.028
Lag−1 max. wind gust [m/s]
0.027
0.00
0.25
0.50
0.75
1.00
Reduction in risk
0.8
0.6
0.4
0.2
b
Random forest (RF)
variable importance ranking of top ten features
Mean pressure [hPa]
0.023
Max. pressure [hPa]
0.022
Min. PT [°C]
0.019
Lag−3 min. dewpoint temp. [°C]
0.011
Min. temperature [°C]
0.0099
Min. pressure [hPa]
0.0094
Lag−2 max. temperature [°C]
0.0092
Lag−1 max. temperature [°C]
0.0091
Lag−7 mean pressure [hPa]
0.0062
Lag−3 mean pressure [hPa]
0.0059
0.00
0.01
0.02
0.03
Permutation−based importance
0.020
0.015
0.010
c
Support vector regressor (SVR)
variable importance ranking of top ten features
Mean pressure [hPa]
0.023
Max. pressure [hPa]
0.022
Min. pressure [hPa]
0.019
Lag−6 wind direction [°]
0.0075
Min. PT [°C]
0.0069
Lag−3 min. pressure [hPa]
0.0064
Sunshine duration [h]
0.0062
Mean vapour pressure [hPa]
0.0061
Mean humidity [%]
0.0059
Mean temperature [°C]
0.0059
0.00
0.01
0.02
0.03
Permutation−based importance
0.020
0.015
0.010

**Supplementary Figure S3. Variable importance of boosted generalized additive model (GAM), random forest (RF), and support vector regressor (SVR).** The bar plots indicate the top ten variables of the fitted **(a)** GAM, **(b)** RF**,** and **(c)** SVR models for daily acute ischemic stroke admissions, respectively. GAM identified maximum and mean pressure as the top two variables, while weekend status was selected as the third most important variable. GAM als included a combination of variables based on wind, temperature and humidity among the top ten variables. RF and SVR identified mean and maximum pressure as the two top variables. Similar to the extreme gradient boosting (XGB) model, RF also revealed minimum perceived temperature (PT) as the third most important variable. SVR selected minimum pressure as the third most important variable. Overall, RF mainly highlighted temperature and pressure-related features in the top ten. In contrast, SVR included a combination of variables based on wind, sunshine, humidity, and temperature, similar to GAM.
